# Changes in eating habits and sedentary behavior during the COVID-19 pandemic in adolescents with chronic conditions

**DOI:** 10.1101/2021.04.16.21255582

**Authors:** Bruna Caruso Mazzolani, Fabiana Infante Smaira, Camilla Astley, Amanda Yuri Iraha, Ana Jessica Pinto, Isabela Gouveia Marques, Milla Cordeiro Amarante, Nathalia Saffioti Rezende, Sofia Mendes Sieczkowska, Tathiane Christine Franco, Luana Cristina do Amaral Miranda, Lívia Lindoso, Alberto Carame Helito, Jane Oba, Ligia Bruni Queiroz, Rosa Maria R Pereira, Hamilton Roschel, Clovis Artur Silva, Bruno Gualano

## Abstract

**Purpose:** To report on the impact of the COVID-19 outbreak on eating habits and sedentary behavior among adolescents with multiple chronic conditions (n=347) from a tertiary, referral hospital *vs*. healthy peers.

**Methods:** This observational study was conducted in Sao Paulo (Brazil) between July and October 2020, period in which a set of social distancing measures to contain the pandemic.

**Results:** The main findings of this study were that adolescents with chronic conditions showed important changes in eating habits (e.g., less often consumption of convenience foods and more often eating in front of television than before quarantine). Also, 86.8% of adolescents with chronic conditions reported increasing screen time during pandemic. No major differences were observed between patients and controls.

**Conclusions:** Adolescents with chronic conditions exposed to pandemic showed substantial changes in lifestyle, stressing the need for specific care to mitigate poor eating habits and excessive sedentary behavior in this group.

## INTRODUCTION

Despite growing evidence showing that pediatric population is less susceptible to severe Coronavirus Disease 2019 (COVID-19)(Kabeerdoss, et al., 2021), adolescents have been refrained from school, sports and social activities. Among healthy adolescents, school closures and home confinement were shown to increase unhealthier eating habits, such as increases in snacking, eating while watching television, and consumption of sweets and fried foods.(Ruiz-Roso, et al., 2020) Social distancing measures also scaled up sedentary behavior in teenagers.(Pecanha, Goessler, Roschel, & Gualano, 2020)

More strict social distancing measures have been specifically implemented among pediatric groups with chronic conditions (*e*.*g*., autoimmune diseases, gastrointestinal, kidney and hepatic conditions), since they are deemed to be at risk for more severe COVID-19.(Evliyaoglu, 2020; Wang, et al., 2020) Because of the pressure in the health system caused by the pandemic, some of these patients were also deprived of in-person medical assistance, resulting in sub-optimal health care delivery and limited recommendations regarding healthy lifestyle during the pandemic. However, it remains unknown to which extent the pandemic has impacted the lifestyle of adolescents with chronic conditions, among whom poorer eating habits and higher sedentariness could lead to even further overall health deterioration (Bar-Or & Rowland, 2004; Dohrn, Welmer, & Hagströmer, 2019).

This study reports on the impact of the COVID-19 pandemic on eating habits and sedentary behavior in a large cohort of adolescents with multiple chronic conditions *vs*. healthy peers.

## METHODS

This is an observational study conducted between July and October 2020, period in which a set of social distancing measures to contain the spread of COVID-19 were in place in Sao Paulo, Brazil. Participants were recruited from the Children’s Institute of Clinical Hospital (School of Medicine of the University of Sao Paulo), the largest tertiary, referral, teaching hospital in Latin America. Adolescent patients (n=347) aged between 10 and 18 years with juvenile rheumatic diseases (juvenile dermatomyositis [n=23]; juvenile idiopathic arthritis [n=83]. and childhood-onset systemic lupus erythematosus [n=43], gastrointestinal and hepatic conditions (celiac disease [n=12], eosinophilic esophagitis [n=23], inflammatory bowel disease [n=44], autoimmune hepatitis [n=28], and liver transplant [n=50]), and kidney conditions (nephrotic syndrome [n=22], chronic kidney disease [n=7] and kidney transplant [n=12]) were included in this study. Additionally, healthy adolescents (n=95), frequency-matched for age and sex, were recruited to serve as controls. All participants completed an online survey on Research Electronic Data Capture^®^ (REDCap^®^) platform, which included questions about: *i)* demographic characteristics (*i*.*e*., age, sex, ethnicity, and educational level); *ii)* changes in eating habits during the pandemic (*i*.*e*., consumption of convenience foods and home-made meals, eating with others, eating in front of television/tablet/cellphone, mindful eating, and cooking); *iii)* changes in sedentary behavior during the pandemic, as assessed by screen time.

The study was approved by the Research Ethics Committee of Clinical Hospital (approval number: 31314220.5.0000.0068). The consent form was signed digitally by all adolescents and their legal guardians before the beginning of the survey.

### Statistical analysis

Descriptive data are presented as mean and 95% confidence interval (95%CI) for continuous variables and absolute and relative frequency (n [%]) for categorical variables. Potential between-group differences for all dependent variables were tested by generalized estimating equations (GEE) models, based on the assumption of a multinomial distribution, cumlogit link function, and an exchangeable working correlation, with group as fixed factor. All GEE models were adjusted for age and sex. Analyses were performed using the statistical package SAS (version 9.4). The level of significance was set at p&#x2264;0.05.

## RESULTS

The mean age of patients and healthy adolescents was 14.3 years (95%CI: 14.0, 14.5) and 14.2 years (95%CI: 13.7, 14.7), respectively (p=0.753). Baseline characteristics were comparable between groups (all p>0.050). Most patients and controls were male (60.0% and 62.4%), Caucasians (60.0% and 51.4%) and attended public school (65.3 and 74.4%). Most adolescents reported changing daily routine after the implementation of social distancing measures (patients: 91.6%; controls: 92.8%; Table 1).

**Table 1.** General characteristics of participants

|  | Patients with chronic conditions (n = 347) | Healthy controls (n = 95) | *p* |
| --- | --- | --- | --- |
| Age | 14.3 (14.0 - 14.5) | 14.2 (13.7 - 14.6) | 0.753 |
| ***Sex*** |  |  |  |
| Male | 217 (62.4) | 57 (60.0) | 0.675 |
| Female | 131 (37.6) | 38 (40.0) |  |
| ***Self-reported ethnicity*** |  |  |  |
| Caucasian | 179 (51.4) | 57 (60.0) | 0.139 |
| Not-Caucasian | 169 (48.6) | 38 (40.0) |  |
| ***Current educational level*** |  |  |  |
| Not studying | 9 (2.6) | 4 (4.2) | 0.784 |
| Elementary school | 207 (59.5) | 56 (59.0) |  |
| High school | 114 (32.8) | 29 (30.5) |  |
| University | 18 (5.2) | 6 (6.3) |  |
| ***Type of school*** |  |  |  |
| Public | 259 (74.4) | 62 (65.3) | 0.078 |
| Private | 89 (25.6) | 33 (34.7) |  |
| ***Changed daily routine during the COVID-19 pandemic*** |  |  |  |
| Yes | 323 (92.8) | 87 (91.6) | 0.684 |
| No | 25 (7.2) | 8 (8.4) |  |

Data presented as mean and 95% confidence interval (95%CI) or absolute and relative frequency (n[%]).

Changes in eating habits in patients and controls during the COVID-19 pandemic are illustrated in Figure 1 (left panel). A small proportion of the adolescents with chronic conditions (7.5%) stated eating convenience foods more often than before quarantine, whereas 44.3% reported a less frequent consumption than before COVID-19 quarantine. The proportion of patients declaring consuming home-made meals more often than before quarantine was 33.8% *vs*. only 2.9% reporting a less often consumption than before quarantine. The percentage of patients reporting eating with others more often than before quarantine was 21.7%, while 14.2% declared doing so less often than before quarantine. While 21.1% of patients reported mindful eating more often than before quarantine, 11% stated doing it less frequently. An expressive percentage of the adolescents with chronic conditions (32.2%) reported eating in front of television more often than before quarantine *vs*. only 17.4% reporting doing so less frequently than before quarantine. More than a third (35.8%) of the patients reported participation in cooking more often than before quarantine *vs*. only 3.5% declaring doing so less frequently.

**Figure 1.**
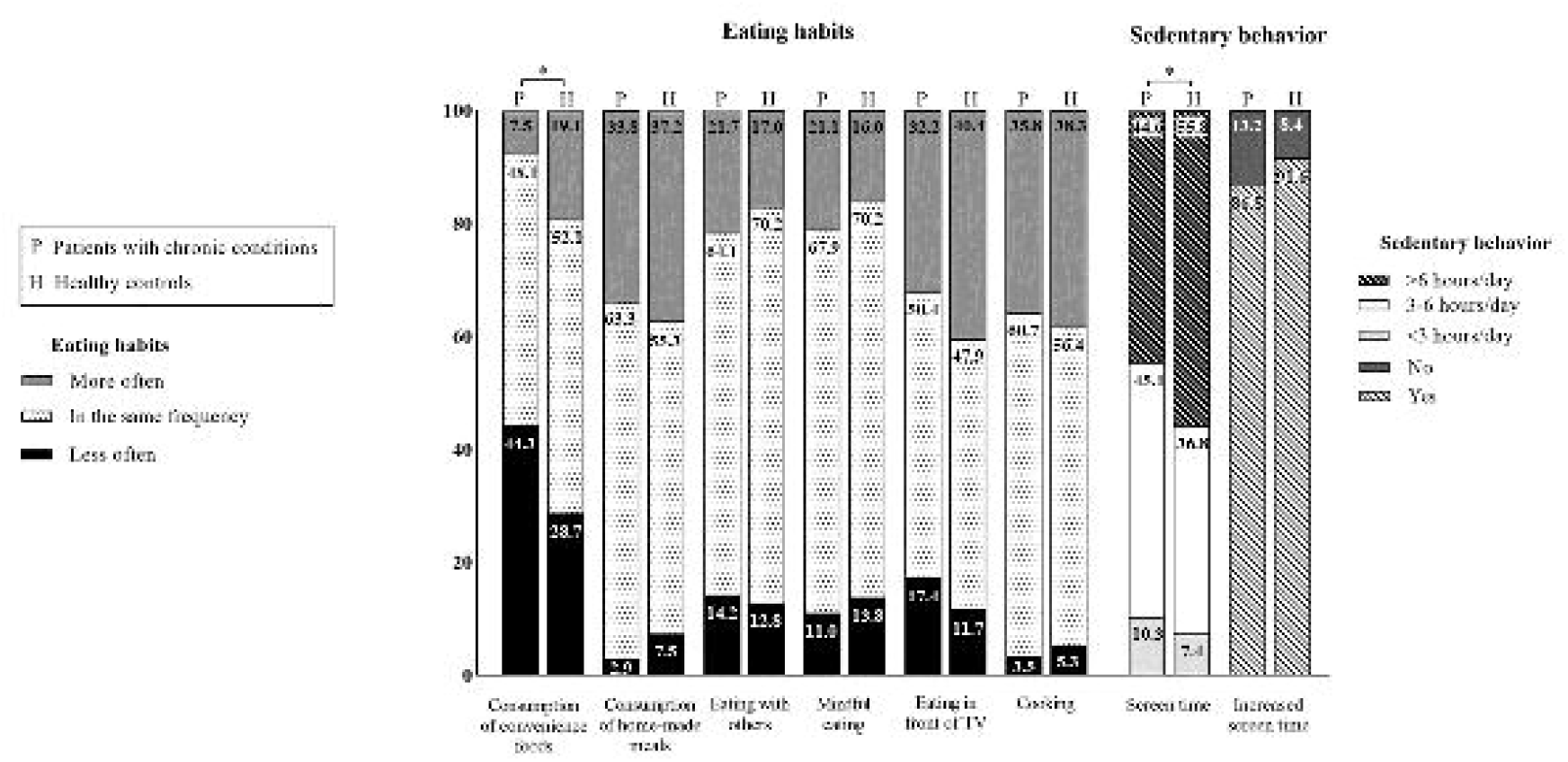
Changes in eating habits and sedentary behavior among adolescents with chronic conditions (n = 347) and healthy controls (n = 95) during the COVID-19 pandemic. *significant difference between groups (p<0.05).

The proportions of patients and healthy controls did not differ for changes in the following eating habits during COVID-19: consumption of home-made meals (p=0.933), eating with others (p=0.612), eating in front of television/tablet/cellphone (p=0.080), mindful eating (p=0.198), and participation in cooking (p=0.827) during the pandemic did not differ between groups. However, the proportion of patients and healthy controls changing consumption of convenience foods was significantly different (more often than before quarantine: 7.5 *vs*. 19.1%; less often than before quarantine: 44.3 *vs*. 28.7%, respectively, p<0.001)

Regarding sedentary behavior (Figure 1, right panel), both adolescents with chronic conditions (86.8%) and their healthy counterparts (91.6%) reported increasing screen time during the pandemic (p=0.219 for between-group comparison). Importantly, a greater part of the healthy adolescents’ sample (55.8%) stated spending more than 6 hours in sedentary behavior when compared with patients with chronic conditions (44.6%), whereas a greater proportion of patients declared spending 3-6 hours (45.1%) and less than 3 hours (10.3%) in sedentary behavior *vs*. healthy controls (36.8 and 7.4%, respectively) (p=0.039).

## DISCUSSION

To our knowledge, this is the first study to report on the impact of COVID-19 pandemic on lifestyle behaviors in a cohort of 347 adolescents with multiple chronic conditions. The main findings of this study were that adolescents with chronic conditions showed important changes in eating habits (*e*.*g*., less often consumption of convenience foods and more often eating in front of television than before quarantine) and substantial increase in screen time, a proxy of sedentary behavior, during the COVID-19 pandemic. Overall, changes in eating habits and sedentary behavior were similar across patients and controls.

The COVID-19 pandemic and the set of social distancing measures adopted to contain disease spread have been shown to impact eating habits worldwide(Ammar, et al., 2020; Ruiz-Roso, et al., 2020) which is also reflected in our sample. Some of these changes appear to be positive, such as the decrease in the frequency of convenience foods consumption and the increase in the proportion of consumption of home-made meals, eating with others, mindful eating and participation in cooking. Conversely, the increased proportion of eating in front of television is worrisome, as it has been associated with unhealth food choices(Avery, Anderson, & McCullough, 2017; Martines, Machado, Neri, Levy, & Rauber, 2019; Trofholz, Tate, Miner, & Berge, 2017) and poor healthy indexes.(Vik, et al., 2013) Considering that parents are generally responsible for providing food, making it available to eat and structuring when and where to eat,(Savage, Fisher, & Birch, 2007) they should be informed on how to mitigate the potentially negative changes in eating habits brought about by COVID-19 pandemic.

Perhaps the most concerning finding of this study was the dramatic increase in the proportion of patients increasing sedentary behavior during the pandemic, with approximately half of the patients reporting screen time over 6 hours/day. While these data align with others(Dunton, Do, & Wang, 2020; Pecanha, et al., 2020) showing excessive sedentary behavior as a consequence of the pandemic, this is of particular relevance for clinical population. Adolescents are known to spend excessive time in sedentary behavior,(Wu, et al., 2017) which may be exacerbated in adolescents with chronic diseases.(Bourdier, et al., 2019; Dohrn, et al., 2019) In this specific group, hypoactivity has been considered a risk factor for poor clinical condition and worse symptoms.(Bar-Or & Rowland, 2004; Gualano, Bonfa, Pereira, & Silva, 2017) Parents and healthcare providers should encourage physical activities and limit screen time whenever possible, particularly in the context of the pandemic. As some chronic conditions (e.g., obesity, immunosuppressive conditions) may aggravate COVID-19 prognosis,(Richardson, et al., 2020; Varley, Ku, & Winthrop, 2021) forcing home confinement, home-based exercise programs may be a valuable and feasible alternative to maintain or increase physical activity levels.(Schwendinger & Pocecco, 2020)

Strengths of this study involve the assessment of a relatively large cohort of patients with several chronic conditions during the most restrictive quarantine in Brazil. This study has limitations, however. First, the cross-sectional design does not allow inferring causative relationships between lifestyle changes and pandemic. Second, the use of self-reported questionnaire may reflect in some degree of imprecision on data reporting. Third, the relatively low number of patients for each disease did not allow for disease-specific sub-analysis. Finally, as a result of the suspension of face-to-face health care in our tertiary hospital (which was adapted to accommodate 1,000 hospital beds exclusively for COVID-19),(Miethke-Morais, et al., 2020) patients’ attendance to in-hospital sessions were not possible, limiting the assessment of more in-depth clinical variables.

In conclusion, adolescents with chronic conditions exposed to social distancing measures due to COVID-19 pandemic showed substantial changes in lifestyle. The findings from this study stress the need for specific care for these patients to mitigate poor eating habits and excessive sedentary behavior.

## Data Availability

Data referred to the manuscript is available under request.

